# The feasibility of targeted test-trace-isolate for the control of SARS-CoV-2 variants

**DOI:** 10.1101/2021.01.11.21249612

**Authors:** William J. Bradshaw, Jonathan H. Huggins, Alun L. Lloyd, Kevin M. Esvelt

## Abstract

The SARS-CoV-2 variant B.1.1.7 reportedly exhibits substantially higher transmission than the ancestral strain and may generate a major surge of cases before vaccines become widely available, while the P.1 and B.1.351 variants may be equally transmissible and also resist vaccines. All three variants can be sensitively detected by RT-PCR due to an otherwise rare del11288-11296 mutation in orf1ab; B.1.1.7 can also be detected using the common TaqPath kit. Testing, contact tracing, and isolation programs overwhelmed by SARS-CoV-2 could slow the spread of the new variants, which are still outnumbered by tracers in most countries. However, past failures and high rates of mistrust may lead health agencies to conclude that tracing is futile, dissuading them from redirecting existing tracers to focus on the new variants. Here we apply a branching-process model to estimate the effectiveness of implementing a variant-focused testing, contact tracing, and isolation strategy with realistic levels of performance. Our model indicates that bidirectional contact tracing can substantially slow the spread of SARS-CoV-2 variants even in regions where a large fraction of the population refuses to cooperate with contact tracers or to abide by quarantine and isolation requests.

---

The frequency of the B.1.1.7 variant of SARS-CoV-2 has grown rapidly from its initial detection in October 2020 to become the dominant strain in southeastern England by 2021. Studies have estimated the new strain is between 40% and 80% more contagious^1,2^. The rapid exponential growth of B.1.1.7, now found in dozens of countries, risks another and potentially higher wave of COVID-19 cases prior to widespread vaccination. Meanwhile, early reports suggest that current vaccines^3^ and prior SARS-CoV-2 exposure^4^ may be less protective against the B.1.351 and P.1 variants now common in South Africa and Brazil.

All three variants share an otherwise rare del11288-11296 (3675-3677 SGF) mutation in orf1ab that can be detected using a single RT-PCR reaction^5^; B.1.1.7 can also be distinguished with the TaqPath diagnostic test^6^, twenty million of which are manufactured weekly^7^. As such, existing COVID-19 testing infrastructure can be used to track the transmission of the new variants. Samples testing positive by other kits can be re-screened^8^ without an emergency use authorization.

Test-trace-isolate (TTI) strategies have been widely used to mitigate the spread of SARS-CoV-2^9^. Models by the present authors^10^ and others^11^ have found that incorporating backwards tracing to identify infector individuals could dramatically increase the efficacy of tracing programs. However, testing delays, mistrust, and low compliance have undermined the confidence of health authorities in the benefits of TTI^12,13^. Moreover, efficacy sharply decreases when caseloads are high^14^, as is true for SARS-CoV-2 – but not yet the variants – in many regions.

Given the current low prevalence of the variants in most jurisdictions and the ability to identify cases of the new variant using existing testing infrastructure, we hypothesised that TTI programs dedicated to controlling them could substantially reduce the harm inflicted prior to widespread vaccination of populations later in 2021, especially if vaccine reformulation is needed. Such programs could be enhanced through incorporation of bidirectional tracing^10^.

However, the effectiveness of TTI strategies varies widely from region to region due to programmatic and population-level differences in variables such as the proportion of cases who share their contact history with contact tracers; the proportion who comply with quarantine and isolation requests; and the overall rate of tracing success. Given this variation, it is unclear whether tracing programs exhibiting realistic levels of performance could feasibly dampen the spread of the new variants.

To evaluate the potential benefits of applying targeted test-trace-isolate to control variants, we applied a branching-process model of COVID-19 contact tracing^10^ to estimate the change in the effective reproduction number achievable across a wide range of parameters.

## Methods

In our branching-process model(Bradshaw et al. 2021), each case generates a number of new cases drawn from a negative binomial distribution according to pre-specified incubation- and generation-time distributions (Table 1). Cases are identified and isolated based on symptoms alone or through contact tracing. Cases either comply with isolation requests or ignore them completely according to some fixed probability of compliance; cases that comply generate no further cases.

**Table 1:** Parameters of the branching-process model.

| Parameter | Value | Sources and Notes |
| --- | --- | --- |
| % asymptomatic carriers | 40% | <sup>17–21</sup> |
| Relative infectiousness of asymptomatic carriers | 45% | Informed by viral loads and tracing results described in <sup>17,21–25</sup> |
| % environmental transmission | 5% | <sup>26,27</sup> |
| Proportion pre-symptomatic transmission | 38% | Informed by <sup>21,22,24,25,28–33</sup> |
| Generation time skew parameter ($\alpha$) | 0.397 | Corresponds to pre-symptomatic transmission rate specified above. |
| % of symptomatic cases identified without tracing | 50% | <sup>34</sup> |
| % of cases who comply with isolation | 50%, 70%, 90% | Assumed |
| Test sensitivity | 70% | <sup>35,36</sup> |
| $R_{\text{base}}$ (before test/trace/isolate) | 1.0 to 2.0 | Assumes a pre-B.1.1.7 $R$ of $\sim 1.0^{1,2}$. |
| Overdispersion | 0.11 | <sup>37</sup> |
| Number of initial cases | 20 | Assumed |
| Incubation period | $6.0 \pm 2.1$ days (lognormal distribution) | <sup>1,38,39</sup> |
| Delay from onset to isolation | $3.8 \pm 2.4$ days (Weibull distribution) | <sup>40</sup> |
| Delay for testing | $1 \pm 0.3$ days (gamma distribution) | Assumed |
| Delay for manual tracing | $1.5 \pm 4.8$ days (lognormal distribution); median 0.5 days | Previous reports suggest most contacts can be traced within one day, but some take longer <sup>41</sup> |

Successful tracing depends on the identified case sharing their contact history with tracers, and on the contact in question taking place within the time window (measured in days pre-symptom onset for symptomatic cases, and days pre-identification for asymptomatic cases). Environmental transmission is assumed untraceable. Symptomatic cases require a positive test before initiating contact tracing.

Each outbreak was initialized with 20 index cases to minimize stochastic extinction and designated as “controlled” if it reached extinction (zero new cases) before reaching 10,000 cumulative cases. Effective reproduction numbers (*R*_eff_) were computed as the mean number of child cases produced per case.

## Results

To investigate the potential for TTI to mitigate the spread of variants, we investigated the effective reproduction number achieved across a range of data-sharing and trace-success rates (Figure 1). To account for uncertainty in variant transmissibility, we explored outcomes for reproduction numbers between 1.2 and 2.0; these values assume that other interventions are already in place.

**Figure 1.**
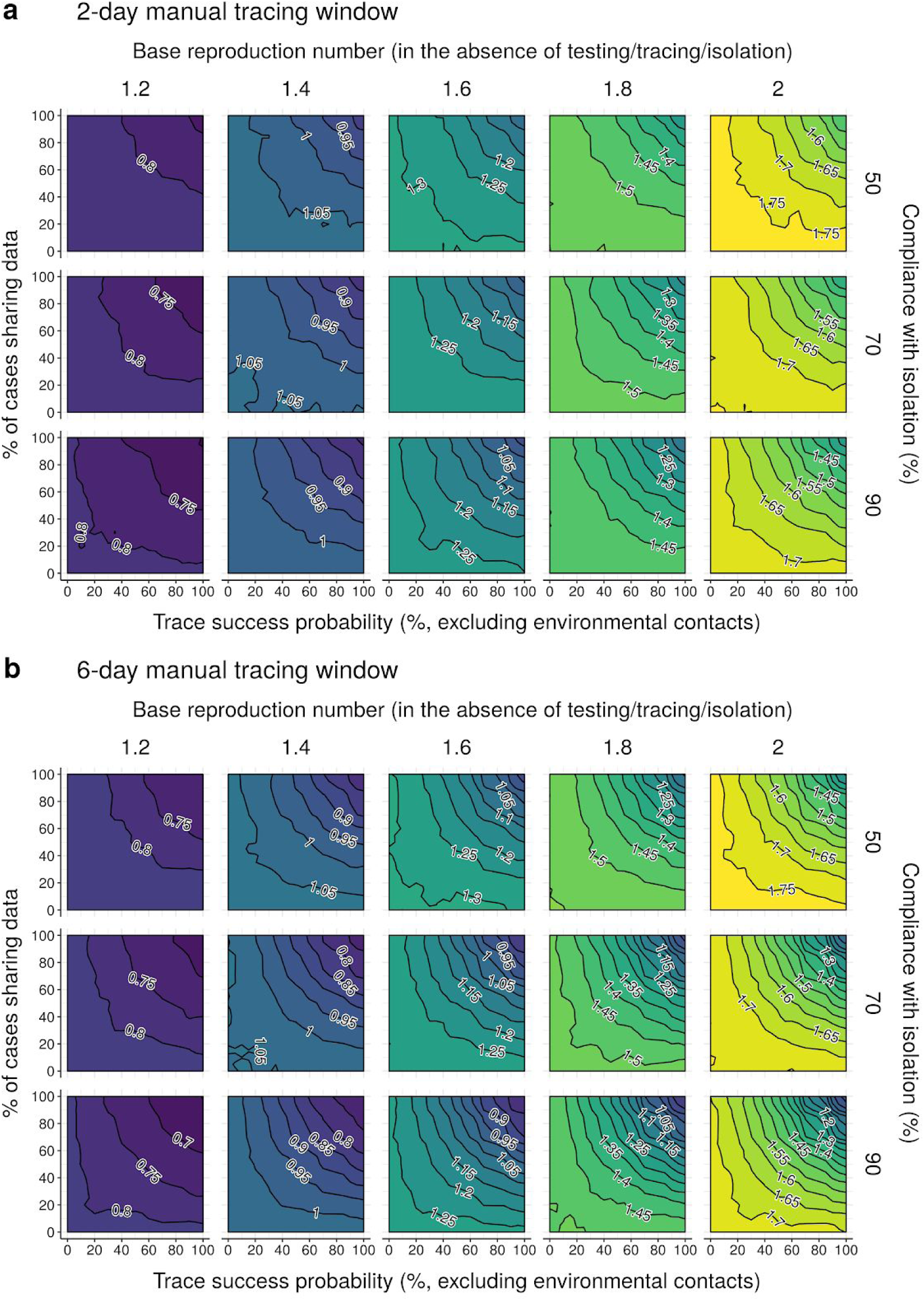
Evaluating the efficacy of bidirectional contact tracing for controlling rare SARS-CoV-2 variants. Neighbour-averaged contour plots, showing *R*eff achieved by bidirectional manual contact tracing with a tracing window of (a) 2 or (b) 6 days pre-symptom onset, under different combinations of trace success probability (x-axis), rate of data sharing with manual contact tracers (y-axis), rate of compliance with isolation and quarantine (row) and base reproduction number (columns). Other disease parameters are specified in Table 1. Isolation of symptomatic cases is sufficient to reduce *R* even when no traces succeed and/or no cases share their data with contact tracers. “Trace success probability” refers to trace attempts that are not otherwise blocked by environmental transmission or refusal to share data.

In the absence of contact tracing, identification and isolation of symptomatic cases alone reduced *R*_eff_ by 0.2 to 0.3 even when quarantine and isolation compliance was low (Figure 1, top rows). When identification and isolation left *R*_eff_ substantially greater than 1 (when base R ≥ 1.4), moderate levels of tracing could have substantial effects.

When contacts were traced up to 2 days prior to symptom onset, ∼60-70% data sharing and trace success rates were required to achieve an *R*_eff_ reduction of at least 0.1, relative to isolation alone. If the window was extended to 6 days pre-onset to enable more effective bidirectional tracing, roughly 45-55% data sharing and trace success was sufficient. Higher levels of data sharing and trace success could achieve substantially larger reductions: in many scenarios, 85% data sharing and trace success reduced *R*_eff_ by >0.2 in the 2-day case and >0.35 in the 6-day case.

Due to the exponential growth of uncontrolled epidemics, small reductions in *R*_eff_ can have a large impact on the total number of downstream cases arising from a given index case over a given timespan. For example, under a simple geometric series approach, reducing *R*_eff_ by 0.1 from a starting value between 1.2 and 2.0 reduces the total number of child cases after 10 generations by 37-43%; an *R*_eff_ reduction of 0.2 results in a reduction in child cases of 61-66%. Given an average generation time of 6 days, 10 generations equates to roughly 2 months – enough time, given sufficient delay in the spread of the new variant, to vaccinate a substantial fraction of the population.

## Discussion

Our results suggest that regions with even moderately functional contact tracing programs focused on the new variants could substantially slow their spread. Given a 2-day window for bidirectionally tracing contacts pre-symptom onset, our model predicts that a program with 70% trace success, 70% data sharing, and 70% compliance with isolation could achieve an *R*_eff_ reduction of at least 0.1 relative to the no-tracing case. Given a 6-day window for efficient bidirectional tracing, regions with just 50% data-sharing, trace success, and isolation compliance could achieve a reduction of 0.1.

Under simple assumptions, such a reduction would reduce the number of child cases produced in two months by roughly 40%, buying time for vaccination to immunise many more people. More effective tracing programs can achieve larger reductions. Higher rates of cooperation might be achieved through home visits by contact tracers; exoneration for anything discovered in the course of contact tracing^13^; and financial and other [support](https://www.masslive.com/coronavirus/2020/08/massachusetts-contact-tracers-can-help-with-ordering-groceries-or-rent-heres-how.html) of people in quarantine and isolation^16^. In principle, concentrating vaccination in communities experiencing out-of-control variant transmission could further impair viral spread and increase the sustainability of TTI.

These results assume a high availability of suitable diagnostic tests and a rapid and consistent testing turnaround. They also take no account of any medical, demographic, geospatial or behavioural variation between cases that could influence the spread of the new variants.

Our results suggest that TTI programs could help slow the spread of more transmissible and vaccine-resistant variants in regions where they are currently rare, providing vital time for widespread vaccination. As TTI efficacy is limited at high caseloads^14^, these findings indicate that tracing programs should immediately prioritise controlling the new variants rather than less transmissible – but currently more widespread – ancestral strains.

## Data Availability

Code Availability: Code for configuring and running the model is publicly available at https://github.com/willbradshaw/covid-bidirectional-tracing

## Acknowledgments

*We thank Aaron Bucher of the COVID-19 HPC Consortium and Amazon Web Services for granting us extra cloud compute credits*.

## Author Contributions

K.M.E. conceived the study. J.H.H. and A.L.L. identified a suitable model framework. W.J.B. designed and programmed the adapted model, advised by the other authors. W.J.B. ran all simulations and generated figures. All authors jointly wrote and edited the manuscript.

## Data Availability

The data supporting the findings of this study are in the main manuscript and the Supplementary Information, and are available at https://github.com/willbradshaw/covid-bidirectional-tracing.

## Code Availability

Code for configuring and running the model is publicly available at https://github.com/willbradshaw/covid-bidirectional-tracing.

## Supplementary Methods

### Structure of the model - Infection dynamics

A new case is infected at some **exposure time**, equal to zero if the case is an index case and otherwise drawn from the **generation time distribution** of its parent case (Table 1). If not asymptomatic, the case develops symptoms at some **onset time** drawn from an **incubation time distribution**. Asymptomatic cases do not develop symptoms, but are still assigned an onset time for the purpose of determining their generation-time distribution.

The number of child cases infected by the case is drawn from a negative binomial distribution, with mean equal to the appropriate reproduction number and heterogeneity determined by the overdispersion parameter *k*. The exposure times of these child cases are drawn from a skewed-normal **generation time distribution** centered on the symptom onset of their parent ^40^, with an SD parameter (ω) of 2 and a skew parameter (α) chosen to give a pre-specified probability of pre-symptomatic transmission (for a symptomatic parent). The generation time distribution for an asymptomatic parent is centered on its “effective” onset time (see above). The shape of the generation-time distribution is the same for all cases.

The expected number of children produced by a case depends on its symptomatic status, and is determined by the overall *R*_0_ value, the proportion of asymptomatic carriers *p*_*asym*_, and the relative infectiousness *x*_*asym*_ of asymptomatic carriers (expressed as a fraction of *R*_0_). Given a reproduction number for asymptomatics of *R*_*asym*_ = *R*_0_ · *x*_*asym*_, the reproduction number of symptomatic cases that produces the desired overall *R*_0_ is given by 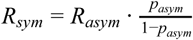.

### Structure of the model - Infection control

Once symptoms develop, a case is **identified** by public health authorities with probability *p*_*isol*_, with the delay from onset to identification drawn from a **delay distribution**. Identified cases are instructed to isolate, and each case complies with probability *p*_*comply*_. Cases that comply generate no further child cases after their time of identification; cases that do not are unaffected. Asymptomatic cases cannot be identified from symptoms, but may be identified via contact tracing from other cases; once identified, they are instructed to isolate as above.

An identified case is **tested**, which takes time drawn from a test time distribution and returns a positive result with probability equal to the sensitivity of the test (since the model does not consider uninfected individuals, the specificity of the test is also not considered). A positive test result is required to initiate contact tracing.

The contacts of cases testing positive are **traced**. Tracing can only proceed outward from a case if they share their contact history with a contact tracer (see below). Tracing can identify the children of the traced case (forward tracing) or its parent (reverse/backward tracing). The speed and success probability of tracing depends on several factors:

- If the contact between the trace originator and the tracee occurred environmentally (determined with probability *p*_*env*_), tracing cannot take place.
- If transmission was not environmental, the contact can be traced manually if:
  - The trace originator shares their contact history with a contact tracer (determined independently for each case with probability *p*_*share*_*manual*_);
  - The time between contact (as above) and the identification time or symptom onset of the trace initiator (whichever came first) is less than the **contact-tracing window** of the tracing system;
  - The tracee is successfully traced by the contact tracer (determined independently for each individual case with probability *p*_*trace*_*manual*_).

Cases that are successfully traced are identified at a time equal to the **trace initiation time** of the trace originator plus a delay time drawn from the appropriate **trace delay distribution**. Identified contacts are quarantined, with an effect identical to isolation and governed by the same compliance variable (i.e. a case either complies with both quarantine and isolation, or neither). Quarantined cases identified through tracing can then be isolated, tested, and traced. If a case is isolated through tracing earlier than they would have been otherwise, child cases whose exposure time would be later than their parent’s new isolation time are eliminated, as are their descendents.

#### Structure of the model - Run initiation

A simulation of an outbreak under the branching-process model is initialised with a given number of index cases (by default 20, in order to reduce the probability of stochastic elimination) and proceeds generation by generation until either no further child cases are generated (extinction) or the run exceeds one of:

1. A **cumulative case limit** of 10,000 cases, or
2. A **time limit** of 52 weeks.

In practice, virtually all runs either went extinct or reached the cumulative case limit; the overall percentage of runs that exceeded the time limit was less than 0.02%, and the highest percentage observed for any single scenario was 1.3%. The cumulative case limit was selected to minimise the chance of a run that would otherwise go extinct being terminated prematurely while preserving computational tractability; in test runs with a cumulative case limit of 100,000 cases, fewer than 2% of extinct runs in any scenario had a cumulative case count of over 10,000.

A terminated run was deemed “controlled” if it reached extinction, and uncontrolled otherwise. The control rate for a scenario was computed as the proportion of runs for that scenario that were controlled. 95% credible intervals on the control rate were computed by beta-binomial conjugacy under a *Beta*(1, 1) uniform prior, as the 2.5th and 97.5th percentiles of the beta distribution *Beta*(1 + *k*, 1 + *n* − *k*), where *n* is the total number of runs for that scenario and *k* is the number of controlled runs. Effective reproduction numbers were computed as the mean number of child cases produced across all cases in a run, averaged across all runs in the scenario. 1,000 runs were performed per scenario.

